# Helmet Use Among E-Bike, Pedal Bike, and E-Scooter Riders in Canberra: Observational and Quasi-Experimental Signage Intervention Study (Phases 1 and 2) [Protocol]

**DOI:** 10.64898/2026.03.04.26347646

**Authors:** Alan Silburn

## Abstract

**Background:** Helmet use is a proven safety measure that reduces the risk of head injury among cyclists and e-scooter riders. Despite legal requirements for pedal bikes and e-bikes in Australia, compliance varies, particularly among users of electric vehicles. The growing popularity of e-bikes and e-scooters in urban areas presents new public health challenges, yet observational data on helmet use, behavioural determinants, and the effectiveness of safety interventions remain limited.

**Aim:** Phases 1 and 2 aim to assess helmet use among e-bike, pedal bike, and e-scooter riders in Canberra, and evaluate the impact of health-benefit and legal-penalty signage on compliance.

**Methods:** This study employs a multi-phase, quasi-experimental observational design across three urban bike paths in Canberra. Phase 1 (Baseline): Helmet use will be recorded via discreet video surveillance, capturing vehicle type, estimated age group, gender presentation, and weather conditions. Phase 2 (Intervention): Two sites will receive signage emphasising either safety benefits or legal penalties, while a third site serves as a control; post-intervention observations will assess changes in helmet compliance.

**Expected Results:** Baseline helmet use is expected to be higher among pedal bike riders than e-bike and e-scooter riders. Signage interventions are anticipated to increase compliance, with potential variation by message type, vehicle type, and rider demographics.

**Trial Registration:** Australian and New Zealand Clinical Trials Registry (ANZCTR) [ACTRN12626000245392]

## 1. SYNOPSIS

Helmet use is a well-established safety measure that significantly reduces the risk of head injury among cyclists and e-scooter riders. Despite Australian legislation mandating helmet use for pedal bikes and e-bikes, compliance remains variable, particularly among users of electric vehicles. The increasing popularity of e-bikes and e-scooters in urban environments presents new challenges for injury prevention. Understanding helmet-wearing behaviour, evaluating the impact of targeted interventions, and linking observational findings to hospital presentations are critical for informing evidence-based public safety policies.

This sub-study forms part of a larger study that employs a multi-method approach comprising three complementary components: a quasi-experimental observational study of helmet use in public settings, retrospective analysis of head injury presentations at The Canberra Hospital, and a cross-sectional survey assessing public attitudes toward helmet use and deterrent fines. Observational data will be collected across three high-traffic urban bike paths, with pre- and post-installation of signage emphasising either health benefits or legal penalties. Hospital data will provide context regarding injury outcomes associated with helmet use trends. Survey data will inform potential policy strategies to increase compliance.

The primary objective of the larger study is to assess helmet use rates among riders of e-bikes, pedal bikes, and e-scooters and to evaluate the effectiveness of signage interventions. Secondary objectives include examining demographic differences in helmet use, exploring associations with head injury presentations, and assessing public perceptions of deterrent fines. This protocol outlines study rationale, design, methodology, ethical considerations, and anticipated outcomes to ensure robust and meaningful findings for injury prevention in Canberra.

## 2. RATIONALE / BACKGROUND

Head injuries associated with cycling and micromobility vehicles represent a substantial public health concern, with consequences ranging from mild concussions to severe traumatic brain injury. Helmet use consistently mitigates the risk of head injury, with protective effects well documented across conventional bicycles, e-bikes, and e-scooters (1,2). For example, a meta-analysis of over 64,000 cyclist crash cases estimated that helmet use is associated with ∼51% lower risk of head injury and ∼69% lower risk of serious head injury compared with non-use, supporting widespread helmet use in cycling safety planning (1). Moreover, Høye’s review suggests that mandatory helmet legislation is associated with reductions of ∼20% in head injuries and ∼55% in serious head injuries (3). In the Australian context, the introduction of universal bicycle helmet laws in the early 1990s was followed by an immediate ∼46% decline in cycling fatality rates, though the effect on nonfatal head injuries is less consistently documented (4).

However, compliance remains inconsistent, particularly among users of electric mobility devices in urban areas. Observational studies in Australia show significant proportions of e-scooter riders not wearing helmets even where mandated (5). A study in Melbourne found that ∼50% of e-scooter injury presentations involved head injuries, while only one-third of riders reported wearing helmets (6). The rapid growth in e-scooter adoption has been mirrored by increasing hospital presentations for related injuries and calls for updated regulatory frameworks (7,8), highlighting the need to explore factors influencing helmet-wearing behaviour.

Previous observational and epidemiological studies suggest that behavioural, infrastructural, and policy factors influence helmet-wearing behaviour. Interventions such as signage emphasising health benefits or legal penalties may promote compliance, though evidence from Australian urban settings is still limited. Complementing observational data with hospital head injury records enables a triangulated view of how helmet behaviour maps to clinical outcomes, while survey data can shed light on the acceptability of enforcement strategies and what might drive behavioural change. In addition, legal context and enforcement levels may further shape compliance patterns.

Helmet laws and fines vary widely across Australian jurisdictions, which may influence helmet-wearing behaviour. In the ACT, riders face a fine of AUD 121 for not wearing a helmet, comparatively modest next to AUD 344 in New South Wales and Tasmania, AUD 227 in Victoria, and AUD 205 in South Australia. Fines are lower in Queensland (AUD 137), Western Australia (AUD 50), and the Northern Territory (AUD 25) (9). Enforcement intensity also differs, with NSW police reportedly issuing ∼500 fines per month (9). These variations suggest that both fine magnitude and policing practices may shape helmet-wearing behaviour, providing critical context for interpreting compliance trends in Canberra and for designing targeted interventions such as signage emphasising health benefits or legal penalties.

Thus, linking helmet use behaviour, signage-based interventions, and hospital head injury trends offers a promising approach to inform effective local injury prevention policy in Canberra.

## 3. AIMS / OBJECTIVES / HYPOTHESES

### 3.1. Aim

Phase 1 aims to quantify helmet use among e-bike, pedal bike, and e-scooter riders in Canberra.

Phase 2 aims evaluate the effectiveness of signage-based public safety interventions in increasing compliance.

### 3.2. Primary Objectives

- Phase 1: To measure baseline helmet use prevalence among riders of e-bikes, pedal bikes, and e-scooters on selected urban bike paths.
- Phase 2: To evaluate changes in helmet use following the implementation of signage interventions emphasising health benefits or legal penalties.

### 3.3. Secondary Objectives

- To examine helmet use patterns by estimated age group, gender presentation, vehicle type, and environmental conditions.
- To identify demographic and contextual predictors of helmet compliance.

### 3.4. Hypotheses

- Compliance will vary by vehicle type, age, and gender presentation
- Helmet use rates will be higher at sites with health-benefit or legal-penalty signage compared with baseline.

## 4. STUDY LOCATIONS AND INTERVENTIONS

Three urban bike paths in Canberra will be selected based on traffic volume, proximity to e-bike and e-scooter rental stations, and prior safety concerns.

Phase 2 Intervention Conditions:

- Control Site: No signage (baseline helmet use).
- Health Benefits Signage Site: Three signs promoting the safety and health benefits of helmet use. Signs will be spaced approximately 50 metres. Imagery on the signs may include (a) an egg wearing a helmet and an egg that has been cracked open; (b) a watermelon wearing a helmet and a watermelon broken; (c) a cartoon head wearing a helmet and a cartoon head with brains exposed.
- Fines Reminder Signage Site: Three signs highlighting legal penalties for non-compliance. Signs will be spaced approximately 50 metres. Signs will have text and may include (a) Not wearing a helmet – Fine = $121; (b) Passenger not wearing a helmet - Fine = $121; (c) Not wearing a helmet – Demerit Points Apply.

Signs will be displayed in accordance with the relevant local authority under the Public Unleased Land Act 2013, section 27 (Movable signs code of practice) (10).

## 5. RESEARCH PLAN / STUDY DESIGN

### 5.1. Data Sources / Collection

#### 5.1.1. Observational Data

Trained research staff will use discreet video cameras to record helmet use at each of the three selected bike paths. Observations will occur over two-week periods during morning and afternoon peaks on both weekdays and weekends to capture typical rider behaviour. Recorded variables will include:

- Helmet use (Yes/No)
- Vehicle type (E-bike, Pedal bike, E-scooter, Pedal scooter)
- Estimated age group (Child, Teen, Adult, Senior)
- Gender presentation (Male, Female, Undeterminate)
- Weather conditions (Fine, Overcast, Rain)

Video footage will be coded using a structured checklist. Observers will remain non-intrusive to minimise behavioural changes due to observation (Hawthorne effect). Data from baseline and post-intervention periods will allow comparison of helmet compliance across sites and time points.

#### 5.1.2. Data Management and Security

All data will be de-identified and assigned unique codes for linkage and analysis. Where possible, video recordings will be stored on secure, password-protected institutional servers accessible only to authorised personnel. Raw video footage will not be shared externally. Data management procedures will comply with institutional, NHMRC, and local privacy guidelines, with routine backups and secure retention for a minimum of five years.

#### 5.1.3. Quality Control and Fidelity Monitoring

Observational coding will be conducted by trained researchers to ensure reliability. Regular review meetings will monitor data integrity, adherence to protocol, and timely resolution of any discrepancies.

### 5.2. Population / Sample Size

#### 5.2.1. Population

Observational data will include all visible riders of e-bikes, pedal bikes, and e-scooters at the selected sites during observation periods, excluding those whose helmet use cannot be reliably determined due to obstruction, darkness, or poor video quality.

#### 5.2.2. Sample Size

Based on prior studies estimating approximately 50% baseline helmet use and an anticipated 10–15% improvement following interventions, a minimum of 400–500 riders per group (approximately 1,200–1,500 total) will be required to achieve 80% power at α = 0.05. Observational periods will capture 30–50 riders per hour per site, requiring approximately 30–40 hours of recording per site.

### 5.3. Statistical Analyses

All statistical analyses will be conducted in R®. Multiple imputation will address missing data where appropriate. Sensitivity analyses will include alternative specifications of period effects, adjustment for baseline covariates, per-protocol analyses limited to high-fidelity clusters, and subgroup analyses by age and baseline compliance.

#### 5.3.1. Observational Data

Descriptive statistics will summarise helmet use prevalence by vehicle type, site, and demographic characteristics. Chi-square tests will be used to compare helmet use across intervention sites and among rider subgroups. Logistic regression models will evaluate predictors of helmet compliance, adjusting for variables such as age, gender presentation, vehicle type, weather conditions, and sign-age intervention.

### 6.4. Limitations

The proposed surveillance windows prioritise commuter peak periods to capture routine transport-related exposure. Recreational and after-hours riding, including intoxication-related use, may therefore be underrepresented. Night-time surveillance was considered but deemed impractical due to safety and feasibility constraints.

## 6. ETHICAL CONSIDERATIONS

### 6.1. Recruitment and Selection of Participants

Observational data will be collected in public spaces without direct interaction.

### 6.2. Inclusion and Exclusion Criteria

Observation includes all riders whose helmet use can be reliably assessed; those obscured or captured in poor-quality video will be excluded.

### 6.3. Limited Disclosure

This study involves limited disclosure to participants, as individual riders will not be directly informed or consented to at the time of observation. This approach is justified because the research involves observation of behaviour in public spaces, does not involve interaction or intervention with individuals, and does not collect identifiable information. Full disclosure prior to observation would likely alter natural behaviour (Hawthorne effect), compromising the scientific validity of the study.

In accordance with National Statement 2.3.1, the research meets the criteria for limited disclosure because: (a) the research carries no more than low risk; (b) the use of limited disclosure is necessary to achieve the research aims; (c) there is no reason to believe participants would object if they were aware of the study; and (d) there is no feasible alternative methodology that would allow collection of valid behavioural data without limited disclosure.

To mitigate ethical concerns, public notification signage will be displayed at observation sites indicating that helmet use is being monitored for research purposes.

### 6.4. Confidentiality and Privacy

All data will be de-identified. Video recordings will be stored securely and accessible only to authorised personnel. Any external sharing for publication or collaboration will be fully anonymised and aggregated.

### 6.5. Data Access and Dissemination

Only study investigators and authorised data managers will access identifiable data (e.g., coded video files). Results will be disseminated at the aggregate level to policymakers, urban planners, cycling advocacy groups, and academic audiences.

### 6.6. Response to Observed Non-Compliance

This study is strictly observational; no interventions will be made with individual riders. Where patterns of unsafe behaviour are identified at the population level (e.g., consistently low helmet use at a site), findings will be communicated to relevant authorities for education or infrastructure improvement.

### 6.7. Legal and Mandatory Reporting Obligations

All procedures comply with applicable privacy and filming laws. Any incidental identification of an immediate risk will be handled in accordance with mandatory reporting guidelines.

### 6.8. Support for Parents and Staff

Participants will receive contact details for the research team and information on support services (e.g., Lifeline, Beyond Blue) if discussions of cycling injuries or fines cause distress.

### 6.9. Aboriginal and Torres Strait Islander Data

The study does not seek to specifically identify Aboriginal or Torres Strait Islander children. Should the research team wish to analyse outcomes by Indigenous status, this will only occur with explicit consent and following approval from an Aboriginal Human Research Ethics Committee (HREC), in line with the NHMRC Ethical Conduct in Research with Aboriginal and Torres Strait Islander Peoples and communities (2018). The protocol will be amended and resubmitted for additional review if such analyses are proposed.

### 6.10. Data Storage and Record Retention

Electronic data will be stored on encrypted institutional servers, with routine backups maintained according to institutional and NHMRC guidelines. All data will be retained for a minimum of five years following study completion, after which secure deletion procedures will be implemented. Hard copies, if generated, will be securely shredded after the retention period. Data management plans will ensure compliance with applicable regulations, including the NSW Health Privacy Manual and NHMRC National Statement on Ethical Conduct in Human Research.

### 7.11. Ethics Approval and Oversight

Ethics approval will be sought from the University of Tasmania HREC before commencement of the relevant study phase. If Indigenous data collection is proposed, additional review will be sought from an Aboriginal HREC. All applications will be submitted and approved before any participant recruitment or data collection begins. The study protocol will be registered in the Australian and New Zealand Clinical Trials Registry (ANZCTR), and the registration details will be publicly available prior to study commencement. Publication of the protocol will also occur before the start of the study to ensure transparency and facilitate reproducibility. Continuous oversight of the study will be maintained throughout its duration, including monitoring participant safety, data integrity, and adherence to the approved protocol. Any adverse events will be promptly documented and reported to the HREC and relevant authorities, and participants will be informed of any developments that may impact their continued participation. Amendments to study procedures will be submitted for ethics review and approval before implementation.

## 7. OUTCOMES AND SIGNIFICANCE

The primary outcome of this study is a detailed characterisation of helmet use patterns among e-bike, pedal bike, and e-scooter riders in Canberra, including variation by vehicle type, rider demographics, and environmental conditions. By systematically observing behaviour before and after the implementation of health-benefit and legal-penalty signage, the study will provide evidence on the effectiveness of targeted public safety messaging in increasing helmet compliance.

The significance of this study is reinforced when considered as part of the larger study. By integrating observational, clinical, and attitudinal data, it will generate a robust, context-specific evidence base for designing interventions aimed at improving helmet compliance among urban riders. Insights from this study may inform policy decisions, including the optimal deployment of educational signage, the calibration of deterrent fines, and broader urban planning strategies to enhance rider safety.

At a population level, the findings have the potential to reduce the incidence and severity of cycling and e-scooter-related head injuries, decrease healthcare utilisation, and promote a culture of safety in urban mobility. The evidence generated will be directly relevant to local policymakers, health authorities, transport planners, and advocacy organisations, supporting the development of scalable, cost-effective, and socially acceptable strategies to enhance helmet use. Ultimately, the study contributes to a broader understanding of behavioural determinants of safety compliance in rapidly evolving urban transport contexts and may serve as a model for similar initiatives in other Australian cities.

## 8. TIMELINES / MILESTONES

- Months 0–2: Site selection, ethics approval, signage design.
- Months 2–3: Baseline observational data collection.
- Months 3–4: Implementation of signage interventions; post-intervention observations.
- Months 4–5: Data cleaning, coding, and analysis.
- Months 5–12: Dissemination of results to stakeholders, manuscript preparation, and policy recommendations.

## 9. PUBLICATION POLICY

Findings will be disseminated via peer-reviewed publications, conference presentations, and reports to policymakers, urban planners, and advocacy organisations. Authorship will adhere to ICMJE guidelines.

**LIST OF INVESTIGATORS AND PARTICIPATING INSTITUTIONS**

**Chief Investigator:**

Dr Brenton Systermans

Course Coordinator & Senior Lecturer, Healthcare in Remote and Extreme Environments, Tasmanian School of Medicine, University of Tasmania.

**Principal Investigator:**

Mr Alan Silburn

Paramedic & Registered Nurse (Division 2), NSW Ambulance / NSW Health

Academic, Western Sydney University Fellow of the Royal Society for Public Health (FRSPH)

**Associate Investigator:**

Dr Sean Chan

Intensive Care Staff Specialist, The Canberra Hospital

State Medical Director, Donate Life ACT

Deputy Director, ACT Trauma Service

**Associate Investigator:**

Dr Thomas Georgeson

Emergency Staff Specialist, The Canberra Hospital

**Participating Institutions:**

University of Tasmania

Churchill Ave, Dynnyrne TAS 7005, Australia

The Canberra Hospital

Yamba Dr, Garran ACT 2605

## Declarations

### Abbreviations

Not applicable

### Human Ethics

Ethics approval has been obtained from the University of Tasmania Human Research Ethics Committee (HREC) on 24^th^ February 2026 [H40540].

### Registration

The Helmet Use in Canberra study has been registered in the Australian and New Zealand Clinical Trials Registry (ANZCTR) [ACTRN12626000245392].

### Consent for publication

The author consents to the publication of this protocol.

### Availability of data and materials

Available at request from the corresponding author.

### Authors’ contributions

The author solely contributed to the conception, design, analysis, and drafting of the manuscript.

## Acknowledgements

Not applicable

